# Numbers of close contacts of individuals infected with SARS-CoV-2 and their association with government intervention strategies

**DOI:** 10.1101/2021.01.20.21250109

**Authors:** Conor G. McAloon, Patrick Wall, Francis Butler, Mary Codd, Eamonn Gormley, Cathal Walsh, Jim Duggan, T. Brendan Murphy, Philip Nolan, Breda Smyth, Katie O’Brien, Conor Teljeur, Martin J. Green, Luke O’Grady, Kieran Culhane, Claire Buckley, Ciara Carroll, Sarah Doyle, Jennifer Martin, Simon J. More

## Abstract

**Background:** Contact tracing is conducted with the primary purpose of interrupting transmission from individuals who are likely to be infectious to others. Secondary analyses of data on the numbers of close contacts of confirmed cases could also: provide an early signal of increases in contact patterns that might precede larger than expected case numbers; evaluate the impact of government interventions on the number of contacts of confirmed cases; or provide data information on contact rates between age cohorts for the purpose of epidemiological modelling.

**Methods:** We analysed data from 140,204 contacts of 39861 cases in Ireland from 1st May to 1st December 2020. Only ‘close’ contacts were included in the analysis. A close contact was defined as any individual who had had > 15 minutes face-to-face (<2 m) contact with a case; any household contact; or any individual sharing a closed space for longer than 2 hours, in any setting.

**Results:** The number of contacts per case was overdispersed, the mean varied considerably over time, and was temporally associated with government interventions. Negative binomial regression models highlighted greater numbers of contacts within specific population demographics, after correcting for temporal associations. Separate segmented regression models of the number of cases over time and the average number of contacts per case indicated that a breakpoint indicating a rapid decrease in the number of contacts per case in October 2020 preceded a breakpoint indicating a reduction in the number of cases by 11 days.

**Discussion:** These data were collected for a specific purpose and therefore any inferences must be made with caution. The data are representative of contact rates of cases, and not of the overall population. However, the data may be a more accurate indicator of the likely degree of onward transmission than might be the case if a random sample of the population were taken. Furthermore, since we analysed only the number of close contacts, the total number of contacts per case would have been higher. Nevertheless, this analysis provides useful information for monitoring the impact of government interventions on the number of contacts; for helping pre-empt increases or decreases in case numbers, and for triangulating assumptions regarding the contact mixing rates between different age cohorts for epidemiological modelling.

## Introduction

Tracing contacts of infected individuals for the purpose of monitoring, isolation or testing is a fundamental pillar of communicable disease control [1]. In the control of COVID-19, contact tracing has been employed since the earliest stages of the pandemic [2] as an essential public health tool to reduce infection transmission, particularly given the high proportion of pre-symptomatic transmission [3].

A national contact tracing service was established in Ireland at the onset of the pandemic with the primary goal of providing advice to contacts of confirmed cases in order to interrupt onward transmission, and to enable targeted testing of those contacts [4]. Whilst these data were collected for this specific reason, the analysis of that data could be useful to inform other aspects of the national pandemic control effort.

Public health interventions are introduced to change the nature of contacts between individuals such that onward transmission is less likely should a contact occur. Examples include the use of face coverings, social distancing or hand washing [5]. In addition, interventions such as restriction of travel or limiting social gatherings may be introduced which are designed to limit the number of contacts of an individual, contributing to a reduction in the overall force of infection for the population. Ultimately, the effectiveness of the overall set of interventions might be determined by indicators of the rate of new infections in the population such as incidence rates (eg 14-day cumulative number of COVID-19 cases per 100,000 [6]) or time-varying reproduction numbers [7]. However, these indicators are by their nature somewhat retrospective and indirect. Analysis of the contacts reported by confirmed cases during the contact tracing process could be used as a more timely metric to evaluate the impact of public health interventions. Furthermore, given that the number of contacts is a key driver of onward transmission, changes in the numeric distribution of contacts per case may provide an early indication of changes in the trajectory of the number of cases over time.

Secondly, the distribution of the number of secondary cases from SARS-CoV-2-infected individuals has been found to be overdispersed [8]. Superspreading individuals could be disproportionately responsible for a high number of secondary infections as a result of a high degree of viral shedding. Alternatively, these individuals might transmit infection to an increased number of susceptible individuals due to the number and nature of their contacts, independent of their degree of viral shedding [9]. Identification of such individuals may aid in targeted interventions towards those most capable of transmitting the virus further.

Finally, the time-varying reproduction number (Rt) is a key metric describing the effectiveness of viral transmission in the population [7]. Current estimates of Rt are primarily inferred from epidemiologic models which involve compartmentalisation of individuals according to their infection and disease status, allowing the number of individuals in the population within the susceptible, exposed, infectious, and recovered (SEIR) compartments to be described at a point in time [10]. Basic versions of this model assume random mixing within the population, such that the contact rates for different cohorts within the population are homogenous. In reality however, the contact rates across different cohorts in the population is not expected to be uniform. If the contact rates between different cohorts in the population is known, this information may be incorporated into SEIR models, and used to estimate age-specific transmission parameters [11,12]. Earlier work has quantified the contact rates of individuals in a ‘natural’ scenario across a range of different countries [13], however no data has yet been published on the impact of the different national-level control measures on contact rates between individuals.

The aim of this study therefore was to use national contact tracing data from Ireland to describe the rates of contact between individuals within and between different age cohorts at different stages of national control measures; determine associations between demographics and contact rates; and evaluate the temporal comparisons between contact distributions and number of cases.

## Methods

### Contact tracing process

Guidelines for public health management of contacts of COVID-19 cases are available [14]. Briefly, contact tracing was conducted at a number of contact tracing centres across Ireland. Following the identification of a confirmed COVID-19 case (the index case), a series of phone calls were triggered:

- The first call informed the index case that they had tested positive and gave them advice on self-isolation. They were asked about their contacts in the 48 h prior to symptom onset or in the 24 hours prior to their test if they were asymptomatic, up until the point they self-isolated.
- The second call collected information about close contacts (see definition below) including name, address, phone number and circumstances of, and date of last contact with case.
- The third call to these contacts informed them of their close contact with a confirmed case and advised them on further action, including referral for testing and restriction of movements.

During the contact tracing process, these data were entered into a Customer Relational Management (**CRM**) database called CovidCare Tracker, which was developed by the HSE Office of the Chief Information Officer in collaboration with the HSE Contact Management Programme. Routine contact tracing was conducted by trained personnel and not public health doctors. However, should the individual conducting the contact tracing consider the case to be one which required the attention of a more trained individual, or a public health doctor, the case was elevated to a status requiring specialist attention, according to the schematic shown in Supplementary Material Figure S1. Similarly, if a case was in a location or situation in which multiple transmission events may have occurred, that situation was flagged as ‘complex’, requiring further detailed investigation by public health doctors. Not all data from cases which were dealt with as ‘complex’ cases were entered into the CRM database (Supplementary Material – Figure S1). Similarly, contacts of Healthcare Workers (**HCWs**) that occurred within the workplace, were dealt with by the hospital occupational health department (or infection prevention and control team) and not entered into the national CRM database, otherwise some HCW workplace contacts were investigated by public health doctors. However, family and social contacts of HCWs were added to the CRM for contact tracing.

### Definition of close contact

Based on European guidance, any of the following were defined as close contacts: any individual who has had > 15 minutes face-to-face (<2 m) contact with a case, in any setting; household contacts (defined as living or sleeping in the same home), individuals in shared accommodation sharing kitchen or bathroom facilities, or sexual partners; healthcare workers, including laboratory workers, who had not worn appropriate PPE or had a breach in PPE during exposure to the case (defined as either direct contact with the case (as defined above), their body fluids or their laboratory specimen, or being present in the same room when an aerosol generating procedure was undertaken on the case); or passengers on an aircraft sitting within two seats (in any direction) of the case, travel companions or persons providing care, and crew members serving in the section of the aircraft where the index case was seated. For those contacts who shared a closed space (including an office or school setting) with a case for >2 hours, a risk assessment was undertaken taking into consideration the size of the room, ventilation and the distance from the case.

A casual contact was defined separately [14]. However, since these contacts were not generally collected from the contact tracing of routine cases, our study only used those contacts that were defined as ‘close’.

### Data management

The CRM data, based on data entries from each call centre, were collected by the Health Service Executive (HSE) under the Medical Officer of Health legislation, collected by the Central Statistics Office (CSO) in compliance with the Statistics Act 1993, pseudonymised, and stored in a centralised database (the CSO C19 Data Research Hub). The CSO C19 Data Research Hub is a secure data repository from which personally identifiable data cannot be exported. These data were accessed through the CSO data hub by the first author for the purpose of this analysis. Access was granted under Section 20(b) of the Statistics Act, 1993, for the purpose of using data collected during the pandemic to aid in the national response. The study was approved by both the National Research Ethics Committee (20-NREC-COV-099) and the Health Research Consent Declaration Committee (20-025-AF1/COV) since the data were used for a purpose other than that for which it was initially collected and since it was not possible to retrospectively obtain consent.

### Initial data cleaning and processing

Data were available in two different datasets. The first (Case data) listed the pseudonymised reference ID of the case, the location of the case (to county level), as well as the DOB rolled back to the first day of each month. Further, an anonymised reference ID was constructed from, and replaced, the DOB and surname of each case and a “current status” column was generated which indicated the status of contact tracing up to the date of the most recent data extract. The second database (Contact data) consisted of the anonymised reference ID of the case, and the following details of each of their reported contacts: DOB (rolled back to the first day of the month), an anonymous reference ID constructed from, and replacing, the DOB and surname of the contact, the type of contact (close, casual, complex, exceptional, other), and the method by which the contact was identified (manually or via a contact tracing mobile phone application - COVID Tracker).

Details of the impact of each data cleaning step on the number of records for analysis are detailed in Supplementary Material Table S1. Briefly, Case 1 data were initially filtered to include only those records where the current status entry indicated that contact tracing had been completed. In addition, duplicate cases were removed by selecting the most recent data entry where multiple entries existed for the same case. Contact data were initially filtered to include only close contacts. Then, contacts identified by COVID Tracker were excluded as these were not linked to a specific case in the contact tracing database. In addition, contacts with no recorded primary case were also removed from the dataset. Finally, we also restricted the analysis to contacts identified after May 1st, due to concerns over the variability of the quality of data collection processes prior to this point.

Case and Contact data were then joined to create an overall dataset at the level of the contact (that is, with each line representing a contact, with a column indicating the reference ID of the primary case). Ages of the cases and contacts were categorised according to age groups corresponding to school-age children, college-age adults, young adults, middle-age adults and retired adults: 0-17; 18-24; 25-39; 40-64 and greater than 65 years of age. Location of the case was recategorised as “Dublin” and “Rest of Country”.

Next, data were collapsed to the case level for analysis (that is, with each line representing a primary case), summarising the overall number of contacts reported by the case (Dataset 1). A second dataset (Dataset 2) was created which summarised the average number of contacts by the age category of the case and the age category of the contact. Histograms of the number of contacts per case showed only a small number of cases with more than 50 close contacts (Supplementary Material Figure S2a and S2b), these records assumed to be erroneous data and were removed from each of the datasets.

### Data analysis

#### Descriptive analysis

The number of contacts per case was summarised using the mean, standard deviation, 2.5^th^, 25^th^, 50^th^, 75^th^ and 97.5^th^ percentiles in the overall dataset, as well as broken down by age of the case and age of the contact.

For each day, the mean number of contacts reported per case was calculated. Next, 7-day rolling averages of the daily mean contacts were calculated for each day. These figures were calculated for the overall dataset, stratified according to the age cohort of the case and stratified according to both the age cohort of the case and the contact.

The timings of key government interventions were extracted from national press releases [15] and noted according to the temporal pattern of contact numbers per case. The start of each defined period of government restrictions was as follows: Stay at Home (27th March); Initial easing (5th May); Phase one easing (18th May); Phase two easing (8th June); Phase 3 easing (29th June); Kildare, Laois Offaly restrictions (7th August); Dublin level 3 (18th September); Donegal level 3 (25th September); National level 3 (6th October); Border counties level 4 (15th October); National level 5 (21st October to 1^st^ December).

#### Model 1 – modelling number of contacts per case

Preliminary analysis demonstrated that the number of contacts per case were overdispersed. Therefore, we chose to model the number of contacts per case using a negative binomial rather than a Poisson regression model. Each time period of the pandemic was coded according to the government intervention level and modelled as a fixed categorical variable. Age of the case was offered to the model as both a categorical and continuous variable. When modelled as a continuous variable, age was modelled using a cubic regression spline. The form of the variable resulting in best model fit as determined by the Akaike Information Criterion (AIC) was used in the final model. Region (Dublin, Rest of Country) and gender were offered to the model as categorical variables.

#### Models 2 and 3 – comparing temporal breakpoints in cases and contacts

To compare temporal breakpoints in the numbers of cases and contacts, second and third negative binomial models were constructed in which day of the year (YDAY) was modelled as a piecewise linear variable. In Model 2, the outcome variable was the number of contacts per case, whereas the outcome variable in Model 3 was the number of cases recorded on that day. For each, the optimal position of the breakpoint was automatically selected using the ‘segmented’ package in R. The number of breakpoints was selected by running separate models varying the number of breakpoints from 2 to 20 and selecting the number with the lowest AIC. The duration in time between points at which number of contacts began to increase or decrease, were compared with points at which the number of cases began to increase or decrease.

#### Assessment of model fit

Model fit was assessed by comparing real versus predicted contact counts according to different subgroups of predicted values (deciles) and according to each month of the pandemic and each age cohort of the cases.

All data manipulation and analyses were conducted in R version 3.3.1 [16], using the “dplyr” [17], “lubridate” [18], ‘mgcv’ [19], ‘segmented’ [20] and “zoo” packages [21] plots were generated using “ggplot2” [22].

## Results

### Descriptive information

Case and Contact datasets consisted of 79212 and 223651 records respectively at initial read in. The impact of different filtering stages is shown in Supplementary Material Table S1. After joining these data, collapsing to the case level and conducting the data cleaning steps outlined in Supplementary Material Table S1, Dataset 1 consisted of 39861 case records. After separating records according to the age cohort of the contact, Dataset 2 consisted of 251565 records.

A histogram of the number of contacts reported per case is shown in Supplementary Material Figure S2. Over the whole time period studied, the mean number of close contacts per case in Dataset 1 was 3.5 contacts, standard deviation was 4.2, whilst the 2.5th, 25^th^, 50^th^, 75^th^ and 97.5th percentiles were 0, 1, 2, 5 and 14 contacts per case respectively.

Mean number of contacts by variable is shown in Table 1. When stratifying by age cohort of the case, the mean and standard deviation of the number of contacts per case was 3.1 and 5.1; 4.5 and 5.0; 3.6 and 4.1; 3.3 and 3.6 and 2.4 and 3.4 contacts per case for the 0-17, 18-24, 25-39, 40-64 and 65 years old and over categories, respectively.

**Table 1.** Descriptive statistics of the mean number of contacts reported per case.

|  |  |  |  | Percentile |  |  |  |  |
| --- | --- | --- | --- | --- | --- | --- | --- | --- |
| Category | n | Mean | S.D. | 2.5th | 25th | 50th | 75th | 97.5th |
| **Age** |  |  |  |  |  |  |  |  |
| 0-17 | 6353 | 3.1 | 5.1 | 0 | 0 | 2 | 4 | 17 |
| 18-24 | 7590 | 4.5 | 5.0 | 0 | 1 | 3 | 6 | 18 |
| 25-39 | 10089 | 3.6 | 4.1 | 0 | 1 | 3 | 5 | 14 |
| 40-64 | 12490 | 3.3 | 3.6 | 0 | 1 | 2 | 4 | 12 |
| >64 | 3339 | 2.4 | 3.4 | 0 | 0 | 1 | 3 | 12 |
| **Gender** |  |  |  |  |  |  |  |  |
| Female | 20424 | 3.6 | 4.3 | 0 | 1 | 3 | 5 | 14 |
| Male | 19415 | 3.5 | 4.3 | 0 | 1 | 2 | 5 | 15 |
| Not reported/neutral | 22 | 2.3 | 2.6 | 0 | 0 | 1 | 4 | 7.5 |
| **Region** |  |  |  |  |  |  |  |  |
| Dublin | 12748 | 3.5 | 4.2 | 0 | 1 | 2 | 5 | 14 |
| Rest of Country | 27113 | 3.5 | 4.4 | 0 | 1 | 2 | 5 | 15 |
| **Intervention period** |  |  |  |  |  |  |  |  |
| Stay at Home | 699 | 2.1 | 1.9 | 0 | 1 | 2 | 3 | 6.6 |
| Initial easing | 1453 | 2.4 | 2.4 | 0 | 1 | 2 | 3 | 8 |
| Phase 1 reopening | 960 | 2.7 | 3.0 | 0 | 0 | 2 | 4 | 10 |
| Phase 2 reopening | 183 | 4.3 | 4.5 | 0 | 1 | 3 | 6.5 | 15.8 |
| Phase 3 reopening | 715 | 5.1 | 5.4 | 0 | 2 | 4 | 6 | 20.0 |
| Kildare, Laois Offaly restrictions | 4645 | 5.4 | 6.6 | 0 | 1 | 3 | 7 | 25 |
| Dublin level 3 | 1848 | 5.0 | 6.1 | 0 | 1 | 3 | 6 | 22 |
| Donegal/Dublin level 3 | 3883 | 4.0 | 4.7 | 0 | 1 | 3 | 6 | 16 |
| National level 3 | 6518 | 3.7 | 4.1 | 0 | 1 | 3 | 5 | 14 |
| Border counties level 4 | 4266 | 2.8 | 3.3 | 0 | 0 | 2 | 4 | 11 |
| National level 5 | 15051 | 2.9 | 3.2 | 0 | 1 | 2 | 4 | 11 |

When stratifying by the age cohort of the contact, cases reported a mean and standard deviation of 0.8 and 2.1; 0.6 and 1.6; 0.7 and 1.4; 0.9 and 1.4; 0.2 and 0.9 contacts who were in the 0-17; 18-24; 25-39; 40-64; ≥64; and unknown or unrecorded age cohorts, respectively.

### Temporal plots

Temporal analysis of the number of contacts reported over time and by region (Figure 1) shows that at the end of the “stay at home” phase of government interventions, the average number of contacts per case was at a minimum of less than 2 contacts per case at the beginning of May. Subsequently, the average number of contacts increased to approximately 6 at the three different time points: beginning of July, beginning of August and midway through September. After this period, the number of contacts dropped to a mean of 4 at the beginning of October, stayed relatively constant for approximately 1 week before dropping to approximately 2.6 at the beginning of Level 5 restrictions. Subsequently the average number of contacts per case began to increase in both Dublin and Rest of the County cases.

**Figure 1.**
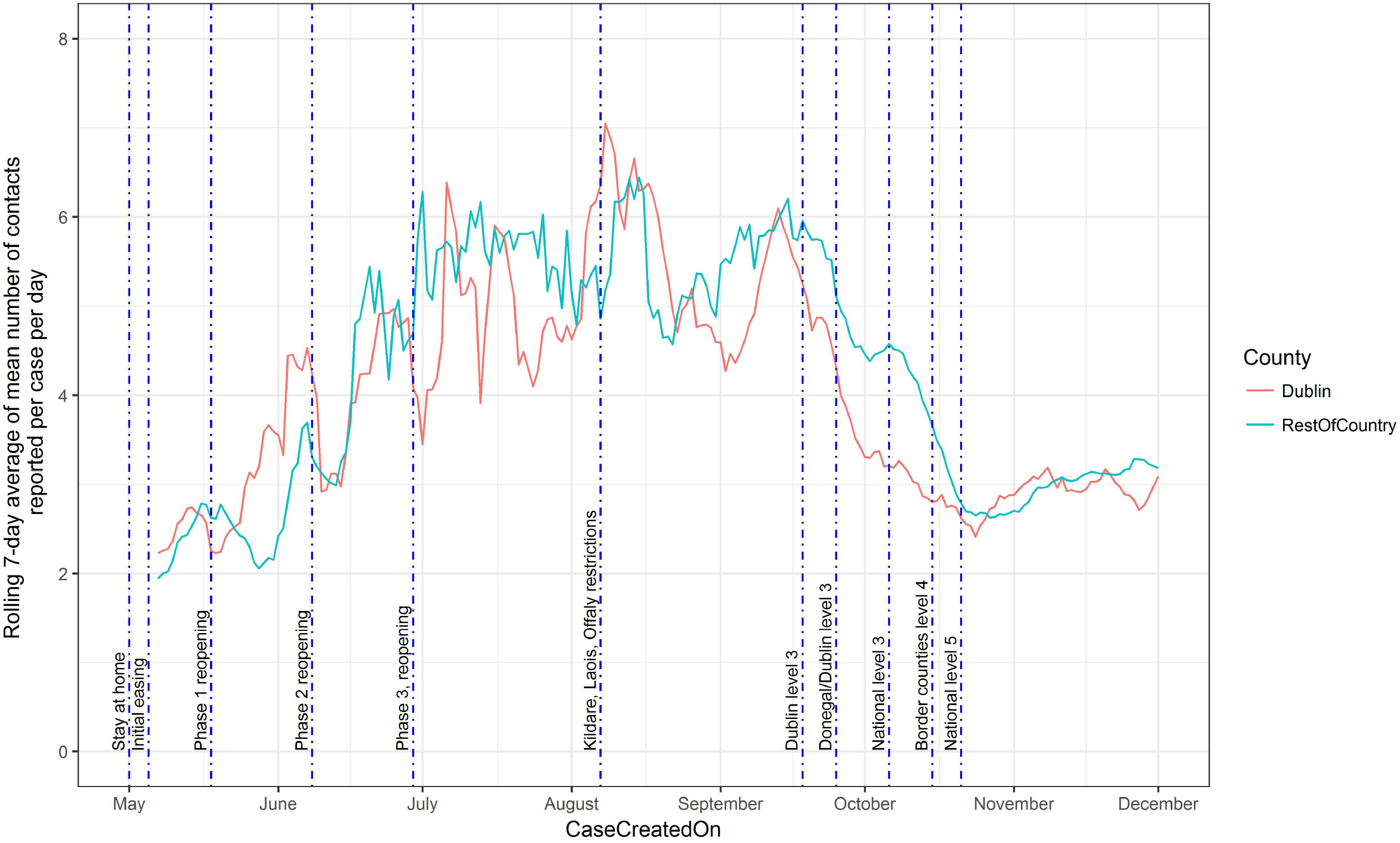
Rolling 7-day average of the mean number of contacts per case per day in Ireland during 2020, by region of the contact (Dublin, Rest of Ireland). The timing of the start of key government restrictions are marked with vertical lines.

Figure 2 shows the temporal pattern in mean number of contacts reported per case according to the age of the case. Overall, the number of contacts are highest in the 18-24 age group. The overall decrease in the number of contacts reported per case that are temporally associated with government restrictions shown from the beginning of October (Figure 1), appear to be largely driven by changes in the 18-24 age cohort (Figure 2).

**Figure 2.**
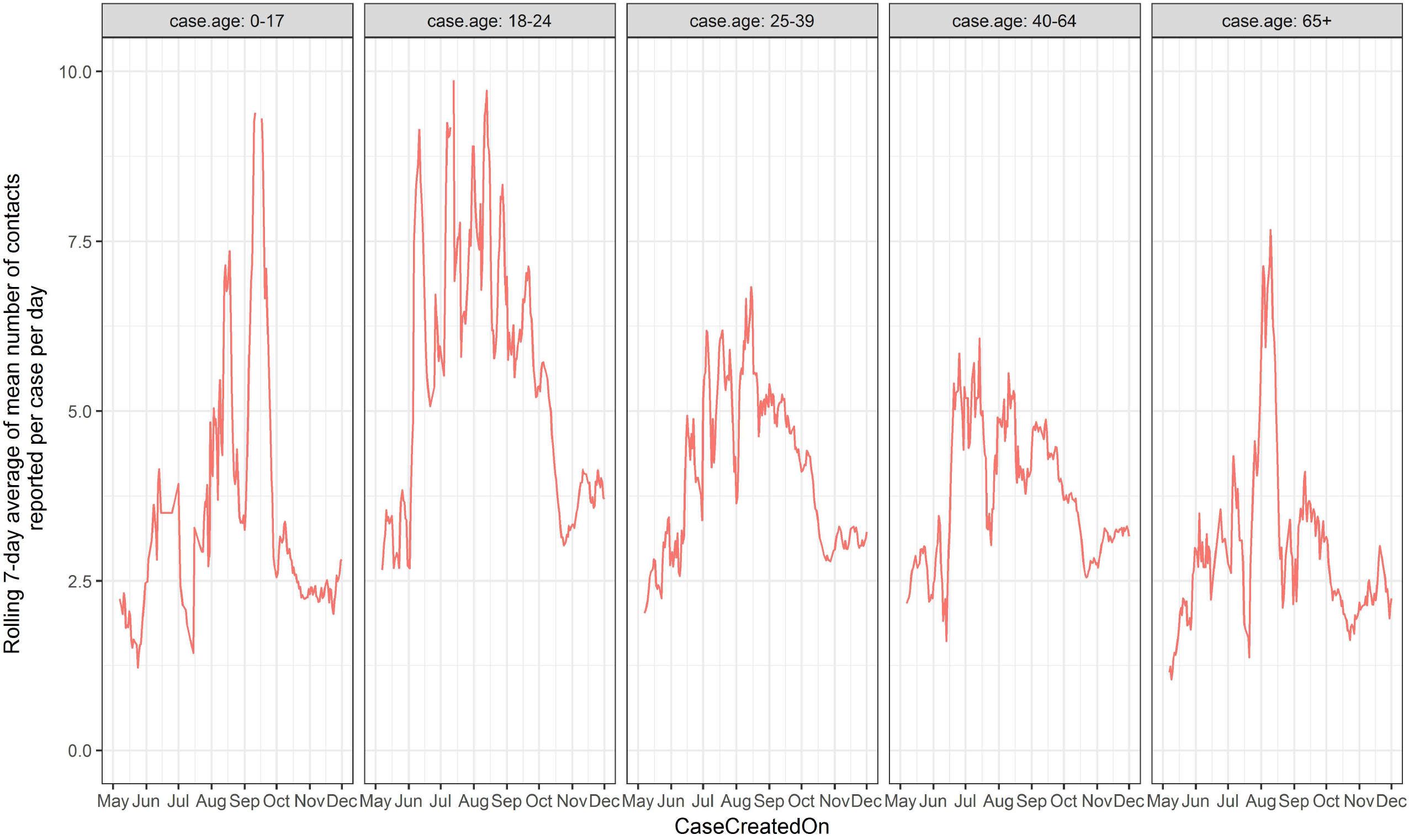
Rolling 7-day average of the mean number of close contacts per day in Ireland Ireland during 2020, according to the age cohort of the case.

Figure 3 shows the temporal pattern in mean number of contacts reported per case according to the age of the case and the age of the contact. This plot shows that the patterns observed in Figure 2 are largely dominated by contact of cases with individuals within the same age cohort.

**Figure 3.**
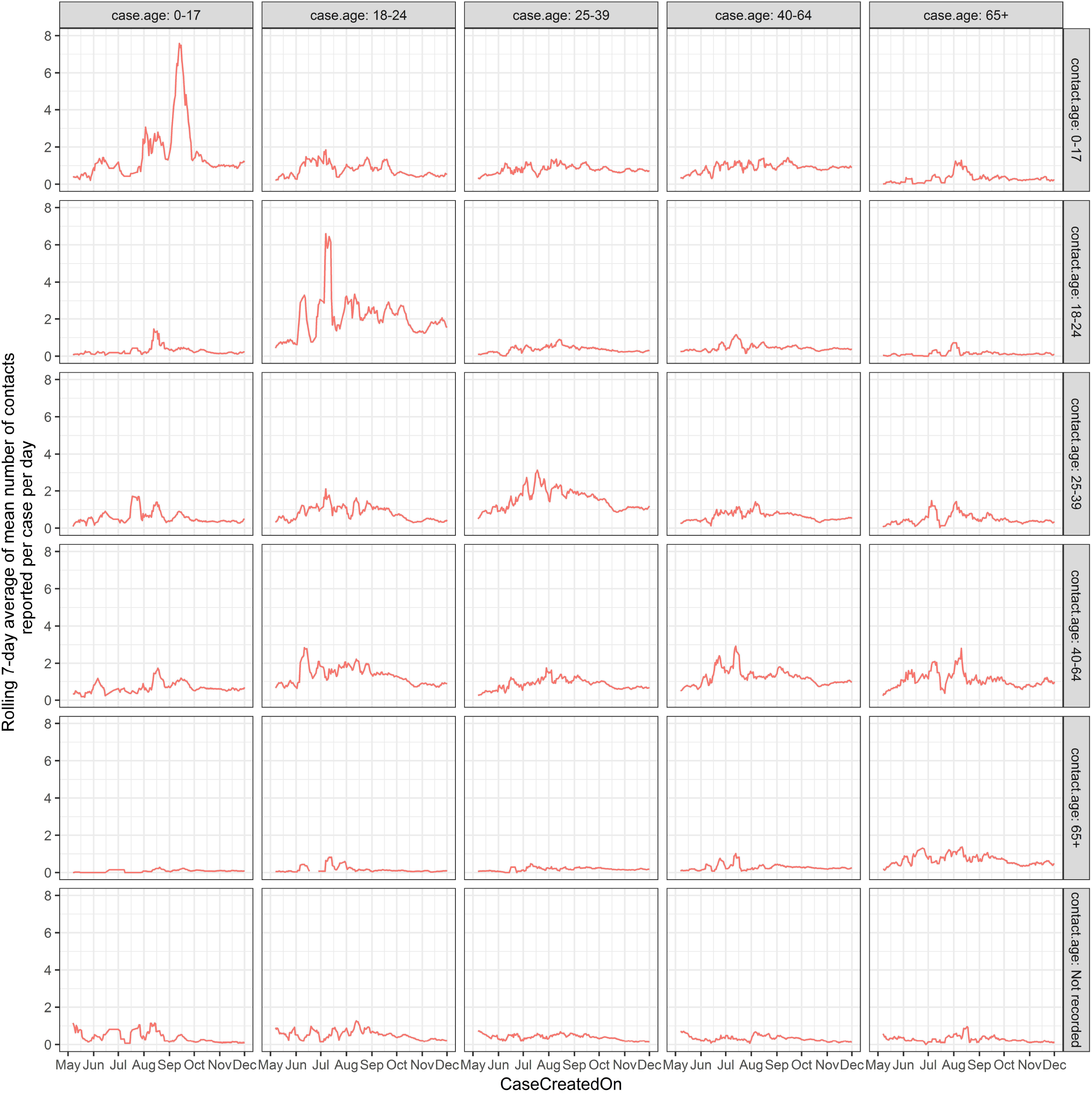
Rolling 7-day average of the mean number of close contacts per day according to the age cohort of both the case and associated contact(s) as reported in Dataset 3.

### Model 1

The results of the negative binomial regression model (Model 1) are shown in Table 2. The Stay-at-home stage of the government intervention was associated with the lowest number of close contacts per case. The log expected count over this time period was 0.9 lower than that of the highest number of contacts per case, Kildare, Laois, Offaly restrictions. The log expected count of contacts was significantly lower in male cases and in Dublin compared with the rest of the country. The best model fit was achieved by modelling age of the contact using a cubic spline, predicted count of contacts per case by age of the case are shown in Figure 4. However, results of second model where age was modelled as a categorical variable are included for illustrative purposes in Table 2. The number of contacts were significantly higher for cases between 18-24 years of age, and lowest in cases in the ≥65 age range. For those in the ≥65 years category, the log expected count of the number of contacts was 0.57 less than those in the 18-25 years category.

**Table 2.**
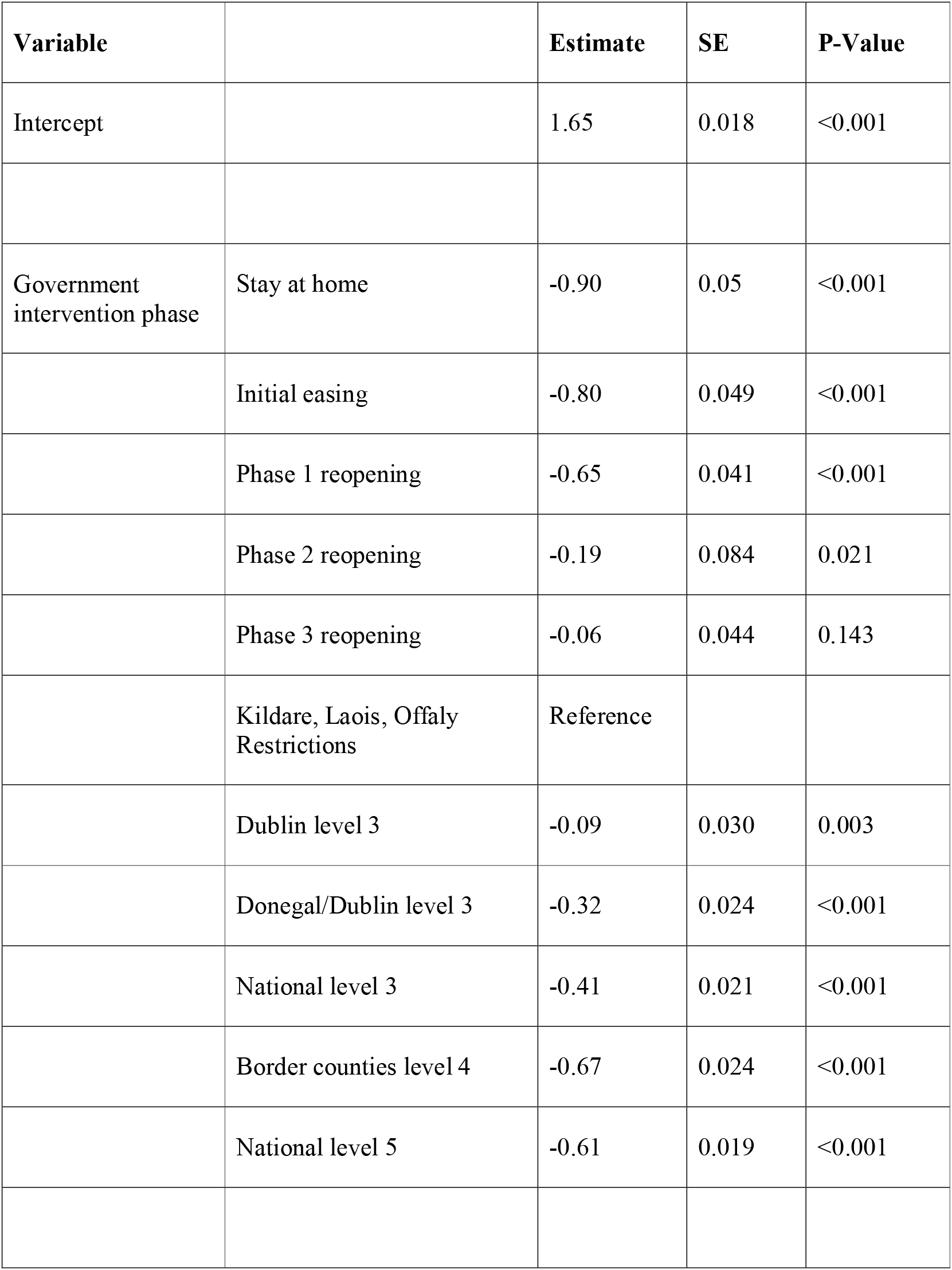
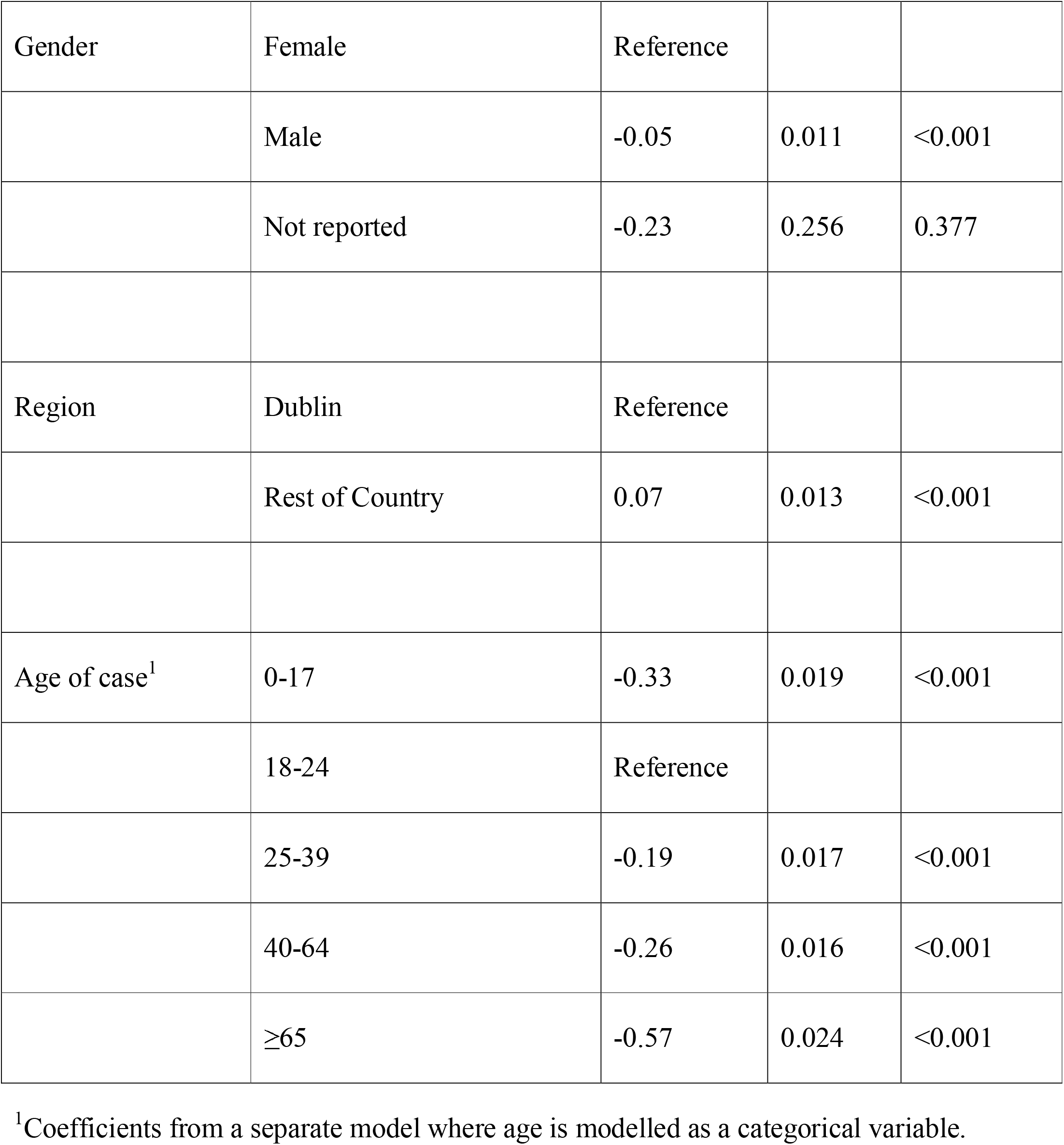
Results of multivariable negative binomial regression model showing association between predictor variables level of government intervention, gender and region and the number of contacts reported per case.

| Variable |  | Estimate | SE | P-Value |
| --- | --- | --- | --- | --- |
| Intercept |  | 1.65 | 0.018 | <0.001 |
| Government intervention phase | Stay at home | -0.90 | 0.05 | <0.001 |
|  | Initial easing | -0.80 | 0.049 | <0.001 |
|  | Phase 1 reopening | -0.65 | 0.041 | <0.001 |
|  | Phase 2 reopening | -0.19 | 0.084 | 0.021 |
|  | Phase 3 reopening | -0.06 | 0.044 | 0.143 |
|  | Kildare, Laois, Offaly Restrictions | Reference |  |  |
|  | Dublin level 3 | -0.09 | 0.030 | 0.003 |
|  | Donegal/Dublin level 3 | -0.32 | 0.024 | <0.001 |
|  | National level 3 | -0.41 | 0.021 | <0.001 |
|  | Border counties level 4 | -0.67 | 0.024 | <0.001 |
|  | National level 5 | -0.61 | 0.019 | <0.001 |
| Gender | Female | Reference |  |  |
|  | Male | -0.05 | 0.011 | <0.001 |
|  | Not reported | -0.23 | 0.256 | 0.377 |
| Region | Dublin | Reference |  |  |
|  | Rest of Country | 0.07 | 0.013 | <0.001 |
| Age of case<sup>1</sup> | 0-17 | -0.33 | 0.019 | <0.001 |
|  | 18-24 | Reference |  |  |
|  | 25-39 | -0.19 | 0.017 | <0.001 |
|  | 40-64 | -0.26 | 0.016 | <0.001 |
|  | ≥65 | -0.57 | 0.024 | <0.001 |

<sup>1</sup>Coefficients from a separate model where age is modelled as a categorical variable.

**Figure 4.**
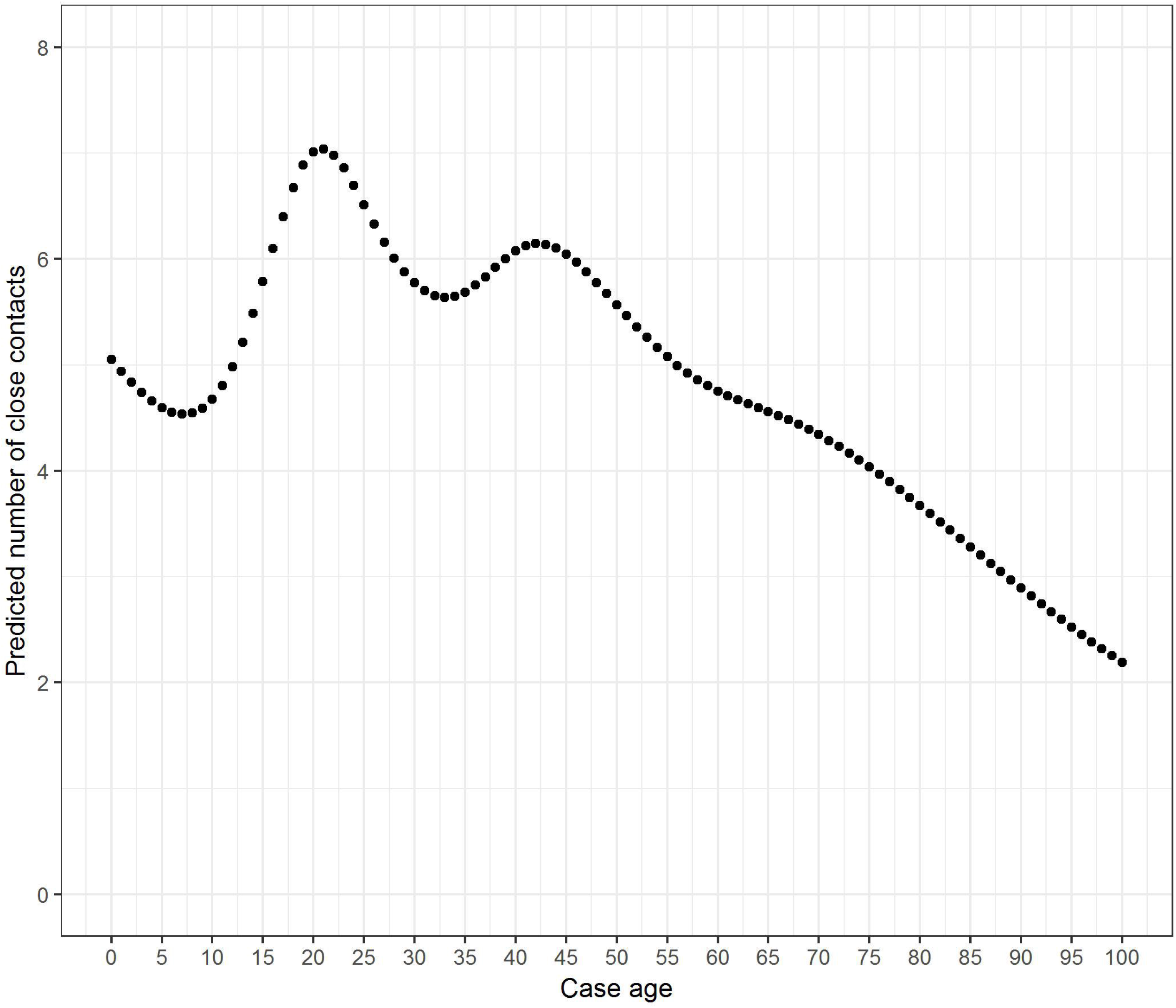
Predicted number of contacts by age of the case when age modelled as a cubic spline.

### Models 2 and 3

Comparison of the temporal breakpoints in contacts and cases is shown in Figure 5. Three breakpoints were identified in the number of contacts per case, whereas two breakpoints were identified in the number of cases per day. Number of contacts per case started increasing from 1st May. A breakpoint indicating an increase in case numbers identified on the 22nd June, approximately 7 weeks later. A breakpoint indicating a decrease in contact numbers was identified on 5^th^ August and a second breakpoint indicating a more rapid decrease in contact numbers on the 7th October, whilst a breakpoint indicating a decrease in case numbers was identified on the 18^th^ October, approximately 9 weeks from the first breakpoint and 11 days after the second breakpoint.

**Figure 5.**
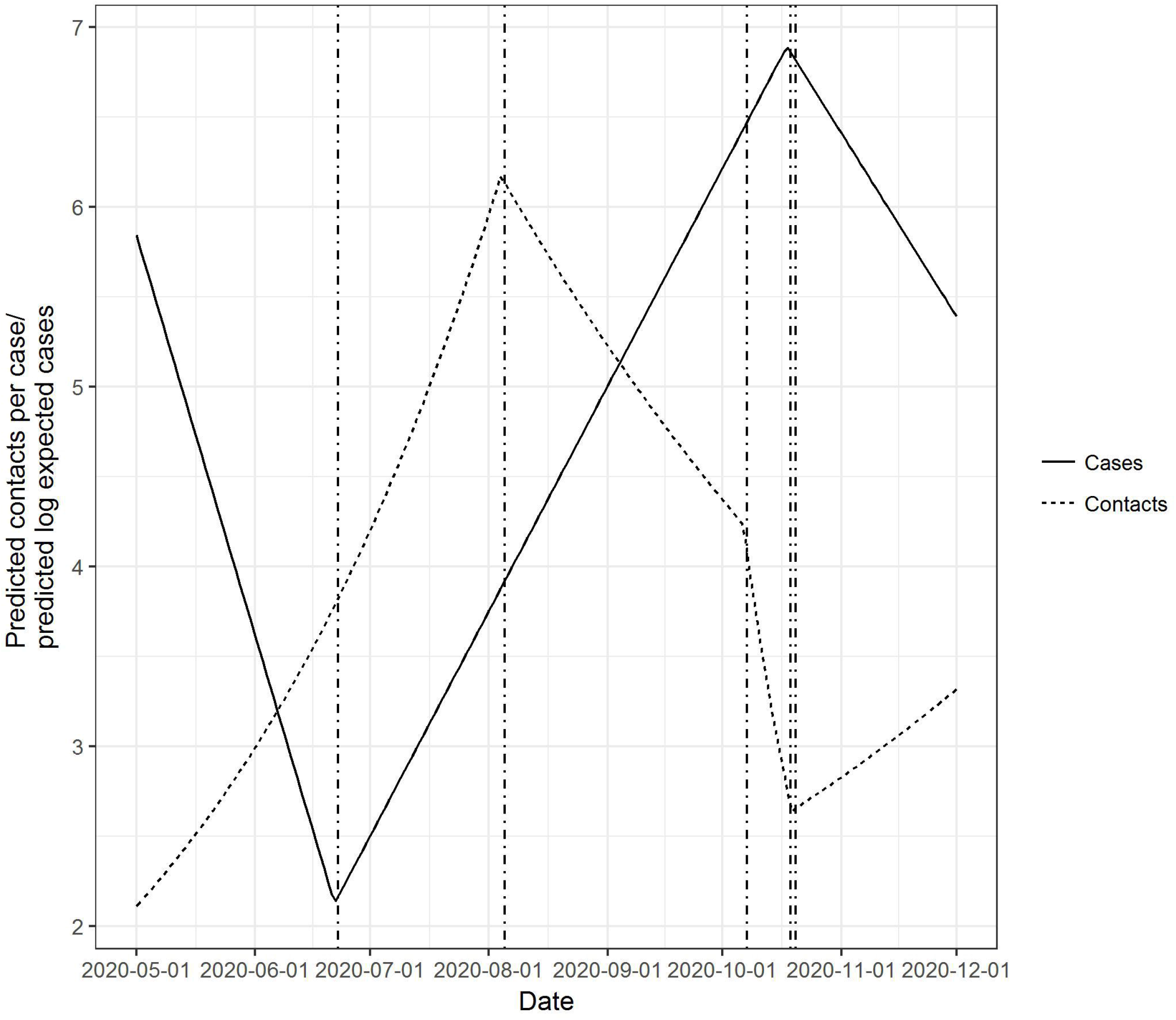
Predicted number of average close contacts and log daily cases. Comparison of breakpoints from segmented regression models.

### Assessment of model fit

Comparing real versus predicted numbers of contacts across each of the predicted deciles indicated a tendency for greater prediction error at greater numbers of contacts. However, the magnitude of this error was relatively small. For example, the root mean squared difference between real and predicted across the first 5 deciles was 0.06, whereas it was 0.22 averaging across deciles 5 to 10. Comparing real versus predicted across each age category demonstrated that the model underpredicted contacts in the 18-24-year-old cohort (mean observed: 4.52, mean predicted: 4.37. Finally, prediction error was also greatest in those months with lower numbers of cases (June to August) with an average error of 0.30.

## Discussion

Contact patterns between infected and susceptible individuals are key drivers of the force of infection for any infectious disease. The temporal association between national interventions and numbers of close contacts of infected individuals may be useful as an early indicator of the effectiveness of interventions. In addition, changes in the pattern of contacts reported from confirmed cases could serve as an early warning for increasing number of cases.

We found that changes in the number of contacts per case were temporally associated with the introduction of government interventions. Such associations must be interpreted with care since there is no possibility for a national control population and there is therefore potential for spurious correlation. However, the regional introduction of interventions facilitates a comparison between regions where different levels of government interventions were introduced (Figure 1). This observation, as well as the closely temporally aligned changes in contacts, suggests that the number of contacts per case were impacted by government interventions. Zhang et al. [23] found a 7-8 fold reduction in overall number of contacts in Wuhan and Shanghai during the social distancing period compared with a baseline survey of contact patterns. No baseline data prior to the pandemic were captured within our population, however, the mean number of contacts per case during the lowest level of government contact restrictions (July to mid-September) was two to three times more than at the highest level of restriction (Stay at home phase, beginning of May), and approximately twice that of the most recent introduction of Level 5 restrictions introduced from the middle of October 2020.

Interestingly, changes in the number of contacts reported per case did not stay constant during the period of restriction. For example, contacts per case reached less than 3 at the point of the introduction of national level 5 restrictions (Figure 1), but immediately started increasing again from this point on. Studying mobility patterns in Germany, Bönisch et al. [24] found similar reductions corresponding to the introduction of government restrictions. However, these authors found that as contacts increased from mid-April, a corresponding increase in case numbers was not observed, which was potentially related to changes in the nature of the contacts between individuals. It is also likely that the nature of contacts change over time as a result of seasonality. For example, a greater proportion of outdoor contacts might be expected in the summer months as opposed to the winter months.

Using segmented regression models, we found that a major breakpoint in the reduction of contacts per case, preceded a decrease in overall case numbers by approximately 9-weeks after the first break point, and approximately 11 days following a breakpoint indicating a rapid decrease in the number of contacts per case. We propose that these delays are likely caused by two factors. Firstly, impacts on case numbers will be delayed by a period corresponding to the latent or incubation period of disease (depending on whether individuals are tested based on symptom onset or not), approximately 4-6 days [25, 26]. Secondly, it is likely that a critical threshold in contacts must be reached before Rt can be forced less than 1. We observed that the number of contacts per case continued to decrease over that 5-week period, and this critical threshold may not have been reached until sometime after the first breakpoint.

The number of contacts per case was overdispersed and we elected to model the number of contacts using negative binomial model, rather than Poisson regression. This finding alone supports the concept that interventions could be targeted towards individuals with very high numbers of contacts. When modelling age as a categorical variable, cases in the 18-24 age cohort had the highest number of contacts overall (Table 2). However, within each age category, the number of contacts was not constant. Consequently, a much better model fit was found when age was modelled as a spline.

A number of limitations must be considered when considering this study. Firstly, it must be remembered that these contact rates are of those that were infected with SARS-CoV-2 at particular points in time; that is, they are not a random sample from the population. Therefore, care is needed in interpretation, in particular when examining temporal associations. For example, at particular points in time in Ireland, older individuals were overrepresented in the numbers of cases. The contacts from these individuals might be expected to be less than in other age cohorts, with many of the most elderly in long term care facilities, and this could be associated temporally with a particular government intervention. As a consequence, changes, for example a reduction, in the average number of contacts could largely be a consequence of changes to case demographics rather than as a result of government restrictions.

However, regional comparison of contact number suggests this is not a significant factor. For example, Figure 1 shows that contacts per case fell much faster in Dublin than the rest of the country following the introduction of regional Level 3 restrictions. Following this, Level 3 was extended nationally, with a corresponding rapid drop in contacts per case across the rest of the country. Furthermore, whilst the number of contacts per case is not representative of the overall population, they are representative of the individuals who were infected at that point in time. Therefore, these data are likely more informative with respect to predicting onward transmission than a random sample from the Irish population might have been, and therefore more likely to be of use in forecasting changes in the trajectory of the disease. Furthermore, these data are gathered from laboratory confirmed cases. Undocumented cases are not included.

Unlike Zhang et al. [23], our study lacks a baseline measurement of contacts prior to the introduction of the disease to the population. However, our observation period is much longer than this earlier analysis, allowing us to compare contact rates over a longer time period including the time of greatest relaxation of government restrictions.

Our data also consists of the overall number of close contacts from infected individuals. A particular definition of a close contact was given for the purpose of contact tracing. In addition, certain complex contact situations (Supplementary Figure S1) were not included in the data collection. These occurrences may relate to situations were much larger numbers of contacts were at risk, therefore the contacts per case available in our dataset may be underestimated. Therefore, the true overall number of contacts is expected to be higher than what we have reported. It is also possible that some cases may not have reported all of their contacts, particularly if they had excessively high numbers of contacts during periods of government restrictions.

In addition, the number of contacts reported in our study represents the total number of contacts in the 48-hour window preceding symptom onset for symptomatic individuals, and the 24-hour period preceding testing for asymptomatic individuals, until the point at which the case self-isolated. For these reasons the absolute number of contacts per case is not comparable between studies. However, the relative reduction in contacts over time may be compared and is useful to monitor changes in contact behaviour over time.

Whilst the contact tracing programme successfully traced the overwhelming majority of contacts, due to a rapid increase in cases late October 2020, approximately 2,000 cases over a 48-hr period were not contact traced. These data are therefore not present in our dataset. However, unless these data were systematically different to the overall population of cases at that time, this short period of incomplete data collection would not have impacted our results.

Finally, we observed an increase in the number of contacts of infected children coinciding with the reopening of schools in September 2020. Whilst an increase in the number of contacts at this time is expected, the process of identifying close contacts in education settings was changed [27], therefore this spike in contacts must be interpreted with caution.

## Conclusions

Analysis of the reported number of contacts per individual in contact tracing data may be a useful early indicator of changes in behaviour in response to, or indeed despite, government restrictions. Whilst these data are representative of cases, and not of the overall population, the data may be a more accurate indicator of the likely degree of onward transmission than might be the case if a random sample of the population were taken.

## Data Availability

Data for this study are not made publicly available

## Acknowledgements

These data were collected as part of the Contact Management Programme (CMP) of the Health Service Executive (HSE). The work of the HSE, and the CMP members in particular, is gratefully acknowledged.

